# Healthcare-associated COVID-19 in England: a national data linkage study

**DOI:** 10.1101/2021.02.16.21251625

**Authors:** Alex Bhattacharya, Simon M Collin, James Stimson, Simon Thelwall, Olisaeloka Nsonwu, Sarah Gerver, Julie Robotham, Mark Wilcox, Susan Hopkins, Russell Hope

## Abstract

**Objectives:** Nosocomial transmission was an important aspect of SARS-CoV-1 and MERS-CoV outbreaks. Healthcare-associated SARS-CoV-2 infection has been reported in single and multi-site hospital-based studies in England, but not nationally.

**Methods:** Admission records for all hospitals in England were linked to SARS-CoV-2 national test data for the period 01/03/2020 to 31/08/2020. Case definitions were: community-onset community-acquired (CO.CA), first positive test (FPT) <14 days pre-admission, up to day 2 of admission; hospital-onset indeterminate healthcare-associated (HO.iHA), FPT on day 3-7; hospital-onset probable healthcare-associated (HO.pHA), FPT on day 8-14; hospital-onset definite healthcare-associated (HO.HA), FPT from day 15 of admission until discharge; community-onset possible healthcare-associated (CO.pHA), FPT ≤14 days post-discharge.

**Results:** One-third (34.4%, 100,859/293,204) of all laboratory-confirmed COVID-19 cases were linked to a hospital record. HO.pHA and HO.HA cases represented 5.3% (15,564/293,204) of all laboratory-confirmed cases and 15.4% (15,564/100,859) of laboratory-confirmed cases among hospital patients. CO.CA and CO.pHA cases represented 86.5% (253,582/293,204) and 5.1% (14,913/293,204) of all laboratory-confirmed cases, respectively.

**Conclusions:** Up to 1 in 6 SARS-CoV-2 infections among hospitalised patients with COVID-19 in England during the first 6 months of the pandemic could be attributed to nosocomial transmission, but these represent less than 1% of the estimated 3 million COVID-19 cases in this period.

## Introduction

Healthcare-associated (nosocomial) transmission was a salient feature of SARS (severe acute respiratory syndrome) and MERS (Middle East respiratory syndrome) outbreaks, with 24% of SARS-CoV-1 infections and 36% of MERS-CoV infections among hospitalised cases (excluding healthcare workers) attributed to healthcare acquisition.^1^ Early in the COVID-19 pandemic, a single-centre study in Wuhan, China, reported that 57 (41%) of 138 COVID-19 cases were nosocomial, of whom 17 were patients already hospitalised for other reasons and 40 were healthcare workers.^2^ High rates of SARS-CoV-2 nosocomial infection among patient-facing healthcare workers and resident-facing social care workers were subsequently reported, in England representing 10% of all COVID-19 cases from 26^th^ April to 7^th^ June 2020.^3^

In the UK, a multi-site study of healthcare-associated COVID-19 during the first two months of the pandemic indicated that 13% of SARS-CoV-2 infections in hospital patients might be nosocomial,^4^ whilst a London hospital reported 15% of COVID-19 cases being hospital-acquired during the same period.^5^ In the context of rapidly increasing case numbers in most European countries during a second phase of the COVID-19 pandemic, and a paucity of available national data from almost all countries, there is a need to quantify healthcare-associated SARS-CoV-2 infection in hospitals. We calculated numbers of community-onset and hospital-onset healthcare-associated laboratory-confirmed COVID-19 cases in England during the first six months of the pandemic by linking national routinely collected data for SARS-CoV-2 test results with hospital admission data.

## Methods

### Data sources and linkage

Public Health England (PHE) collects data on all SARS-CoV-2 (COVID-19) PCR tests from laboratories across England.^6^ Laboratory data systems feed automatically into PHE’s Second Generation Surveillance System (SGSS).^7^ In SGSS, the date the test sample was taken is recorded as the ‘specimen date’. We used this date in our analysis for positive SARS-CoV-2 tests, referred to throughout this paper as the ‘test date’. In cases with multiple SARS-CoV-2 positive tests, the earliest positive test date was retained.

Data on all hospital attendances and admissions in England are collated by NHS Digital and sent daily to PHE via the Secondary Uses Service (SUS) and Emergency Care Dataset (ECDS) data collections for admitted patient stays and Accident and Emergency (A&E) attendances, respectively. SUS data are reported monthly, ECDS daily, both on a mandatory schedule. SUS data are presented in consultant episodes, where a patient is under the continuous care of a single consultant.^8^ Episodes were grouped into spells, with a continuous inpatient (CIP) spell comprising one or more consultant episodes within a single hospital provider. The standard NHS Digital methodology for creating CIPs was adapted to restrict hospital spells to a single provider.^9^ When CIPs overlapped in time within a single provider, they were joined. Hospital records from ECDS and SUS were joined into a single continuous record of patient stay when an A&E attendance ended with a discharge coded as an inpatient admission to the same hospital provider for the same patient. Charlson comorbidity indices were calculated from ICD-10 codes for a spell using the method of Quan *et al*. and grouped as 0, 1 or ≥2 comorbidities.^10^

Mortality data were obtained from the PHE National Incident Coordination Centre (NICC) Epidemiology Cell (EpiCell). These data are derived from four sources: deaths notified by hospitals to NHS England; deaths notified to local PHE Health Protection Teams; laboratory reports where a laboratory-confirmed test result has been linked to a hospital-recorded death; and UK Office for National Statistics (ONS) death registrations.^11^ A COVID-19 death was defined as a death that occurred ≤28 days after the first positive SARS-CoV-2 test.

Hospital records from ECDS and/or SUS were linked to SGSS COVID-19 positive test records deterministically using patient NHS number and date of birth else local hospital patient identifier (hospital number) and date of birth. SUS, ECDS and SGSS extracts were obtained on 09/12/2020, mortality data on 14/10/2020. The study period was bounded by first positive test dates between 01/03/2020 and 31/08/2020.

### Community-onset and hospital-onset classifications

Allocation to a community-onset or hospital-onset category was determined using the first positive SARS-CoV-2 test result paired with hospital record start date (emergency care attendance date or inpatient date of admission, where date of admission is day 1) or end date (inpatient date of discharge) according to the following classifications (illustrated by examples in Figure 1):^12^

**Figure 1:**
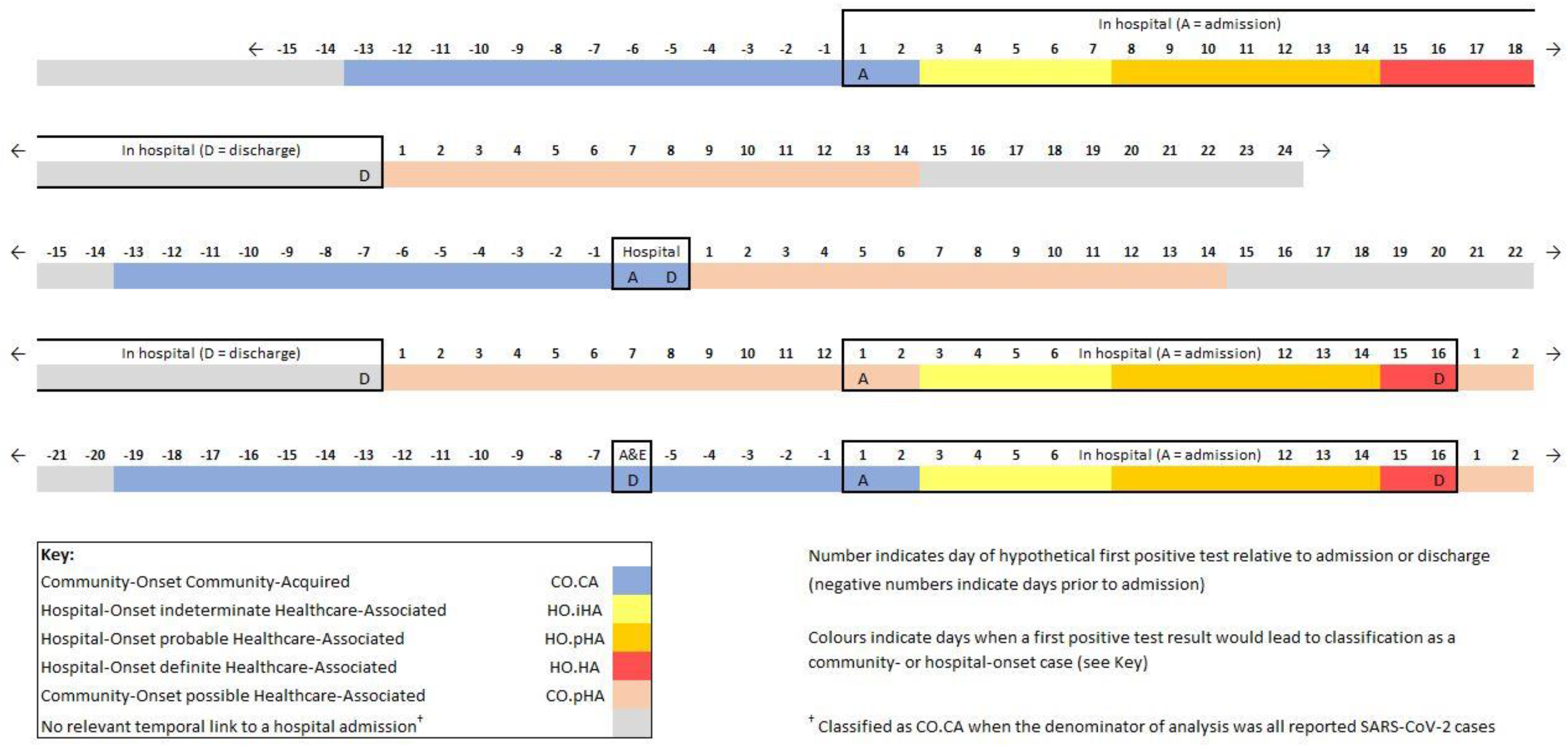
Classification of patients admitted to hospitals in England who tested positive for SARS-CoV-2 as hospital-onset indeterminate, probable and definite healthcare-associated (HO.iHA, HO.pHA, HO.HA), community-onset community-acquired (CO.CA) and community-onset possible healthcare-associated (CO.pHA)

- Community-onset community-acquired (CO.CA): positive test date <14 days pre-admission/attendance and up to day 2 of admission; no prior discharge within 14 days of admission/attendance
- Community-onset possible healthcare-associated (CO.pHA): positive test date ≤14 days post-discharge; if readmitted during this period, up to day 2 of admission where date of readmission is day 1
- Hospital-onset indeterminate healthcare-associated (HO.iHA): positive test from day 3 to day 7 of admission, inclusively
- Hospital-onset probable healthcare-associated (HO.pHA): positive test from day 8 to day 14 of admission, inclusively
- Hospital-onset definite healthcare-associated (HO.HA): positive test from day 15 of admission until day of discharge, inclusively
- Unclassified: All cases which do not meet one of the above criteria, i.e., the positive test did not have a relevant temporal link to a hospital admission or A&E attendance

For each positive test, a single hospital admission was retained for the final onset categorisation. When a patient had multiple hospital admissions, prioritisation was given to an admission overlapping with a positive sample date; when admissions conflicted on the same day in two different trusts, SUS took priority over ECDS data; when a patient had a positive sample between two hospital stays, the completed hospital stay following the positive test was used unless the time between the discharge and positive test was greater than 14 days in which case the admission prior to the test was used. When the only evidence of an admission was an A&E discharge coded as an admission or transfer, the patient was assumed to be still in hospital at the time of data extraction, up to a maximum of 90 days between admission and positive test result, after which, the temporal link is discarded, and the specimen is considered unlinked.

Analyses of community- and hospital-onset COVID-19 excluded SARS-CoV-2 positive cases who had no ECDS or SUS record meeting the temporal criteria for community- or hospital-onset infection (or no ECDS or SUS record) or which were missing both NHS and hospital numbers, except when the denominator for analysis was all reported SARS-CoV-2 positive cases, for which ‘unlinked’ cases were classified as CO.CA.

Length of stay was calculated as the total time (in days) between attendance and discharge, starting with overnight bed-days in A&E, if applicable, or an inpatient admission. For hospital-onset cases, post-test length of stay was calculated as the time (in days) between the first positive test date and the date of discharge. Length of stay for CO.pHA cases are classified as 0 unless the case definition for both CO.pHA and CO.CA are met for the inpatient stay.

Statistical analyses were descriptive, comprising frequencies and percentages for community-onset and hospital-onset classifications stratified by month, region, and provider type (and, for NHS acute and mental health and learning disability trusts, by age group, sex, ethnicity, and Charlson score) and mortality, and median with interquartile range (IQR) for age and length of hospital stay.

#### Ethics

All data were collected within statutory approvals granted to Public Health England for infectious disease surveillance and control. Information was held securely and in accordance with the Data Protection Act 2018 and Caldicott guidelines.

## Results

### Community-onset and hospital-onset COVID-19 cases in England, March-August

Of the 293,204 laboratory-confirmed COVID-19 cases in England with a first positive SARS-CoV-2 test date between March 1^st^ and August 31^st^, 2020, 100,859 (34.4%) were linked to a time-relevant emergency care attendance and/or hospital admission, 167,467 (57.1%) had no time-relevant hospital record and 24,878 (8.5%) had missing NHS and hospital numbers. The proportion of all laboratory-confirmed cases linked to a hospital record declined from a maximum of 79.2% (25,874/32,682) in March to 6.9% (2,054/29,807) in August (Figure S1).

Probable and definite (HO.pHA and HO.HA) hospital-onset healthcare-associated cases represented 5.3% (15,564/293,204) of all laboratory-confirmed COVID-19 cases and 15.4% (15,564/100,859) of cases among hospital patients in England (**Table 1**). Community-onset cases (CO.CA and CO.pHA) represented 91.6% (268,495/293,204) of all laboratory-confirmed cases and 75.5% (76,150/100,859) of laboratory-confirmed cases with a hospital admission; of the latter, 19.6% (14,913/76,150) were possibly healthcare-associated, representing 5.1% of all laboratory-confirmed COVID-19 cases.

**Table 1:** Monthly community-onset and hospital-onset COVID-19 as proportions of all laboratory-confirmed COVID-19 cases and as proportions of all hospital patients who tested positive for SARS-CoV-2.

|  |  |  | Community-onset<br>community-acquired<sup>†</sup> | Hospital-onset<br>indeterminate<br>healthcare-associated | Hospital-onset<br>probable<br>healthcare-associated | Hospital-onset<br>definite<br>healthcare-associated | Community-onset<br>possible<br>healthcare-associated |
| --- | --- | --- | --- | --- | --- | --- | --- |
| **March** | Positive for SARS-CoV-2 |  |  | n=2,583 | n=1,397 | n=2,643 | n=3,380 |
|  | as % of all laboratory-confirmed COVID-19 cases | (N=32,682) | 69.4% (n=22,679) | 7.9% | 4.3% | 8.1% | 10.3% |
|  | as % of hospital patients with COVID-19 | (N=25,874) | 61.3% (n=15,871) | 10.0% | 5.4% | 10.2% | 13.1% |
| **April** | Positive for SARS-CoV-2 |  |  | n=4,570 | n=3,036 | n=3,627 | n=7,216 |
|  | as % of all laboratory-confirmed COVID-19 cases | (N=117,915) | 84.4% (n=99,466) | 3.9% | 2.6% | 3.1% | 6.1% |
|  | as % of hospital patients with COVID-19 | (N=50,119) | 63.2% (n=31,670) | 9.1% | 6.1% | 7.2% | 14.4% |
| **May** | Positive for SARS-CoV-2 |  |  | n=1,254 | n=1,667 | n=1,455 | n=2,700 |
|  | as % of all laboratory-confirmed COVID-19 cases | (N=67,967) | 89.6% (n=60,891) | 1.9% | 2.5% | 2.1% | 4.0% |
|  | as % of hospital patients with COVID-19 | (N=14,905) | 52.5% (n=7,829) | 8.4% | 11.2% | 9.8% | 18.1% |
| **June** | Positive for SARS-CoV-2 |  |  | n=449 | n=597 | n=626 | n=1,035 |
|  | as % of all laboratory-confirmed COVID-19 cases | (N=25,496) | 89.4% (n=22,789) | 1.8% | 2.3% | 2.5% | 4.1% |
|  | as % of hospital patients with COVID-19 | (N=5,590) | 51.6% (n=2,883) | 8.0% | 10.7% | 11.2% | 18.5% |
| **July** | Positive for SARS-CoV-2 |  |  | n=187 | n=179 | n=190 | n=336 |
|  | as % of all laboratory-confirmed COVID-19 cases | (N=19,337) | 95.4% (n=18,445) | 1.0% | 0.9% | 1.0% | 1.7% |
|  | as % of hospital patients with COVID-19 | (N=2,317) | 61.5% (n=1,425) | 8.1% | 7.7% | 8.2% | 14.5% |
| **August** | Positive for SARS-CoV-2 |  |  | n=102 | n=64 | n=83 | n=246 |
|  | as % of all laboratory-confirmed COVID-19 cases | (N=29,807) | 98.3% (n=29,312) | 0.3% | 0.2% | 0.3% | 0.8% |
|  | as % of hospital patients with COVID-19 | (N=2,054) | 75.9% (n=1,559) | 5.0% | 3.1% | 4.0% | 12.0% |
| **Overall** |  |  |  | n=9,145 | n=6,940 | n=8,624 | n=14,913 |
|  | as % of all laboratory-confirmed COVID-19 cases | (N=293,204) | 86.5% (n=253,582) | 3.1% | 2.4% | 2.9% | 5.1% |
|  | as % of hospital patients with COVID-19 | (N=100,859) | 60.7% (n=61,237) | 9.1% | 6.9% | 8.6% | 14.8% |

<sup>†</sup> Denominator = all laboratory-confirmed cases, community-onset includes all COVID-19 cases not linked to a hospital record; denominator = hospital patients, community-onset excludes patients not linked to a hospital record

As monthly proportions of hospital patients with COVID-19, HO.pHA and HO.HA hospital-onset healthcare-associated cases peaked during May and June, at 21.0% (3,122/14,905) and 21.9% (1,223/5,590), respectively (**Figure 2, Figure 3**). The peak in HO.pHA and HO.HA cases occurred in week 22 (27^th^ May to 2^nd^ June) at 26.5% (558/2,109), double the proportion in week 14 (1^st^ to 7^th^ April) at 12.7% (2,342/18,687), which was the week with highest number of laboratory-confirmed cases linked to a hospital record (18,687 linked cases, from a total of 27,671 laboratory-confirmed cases).

**Figure 2:**
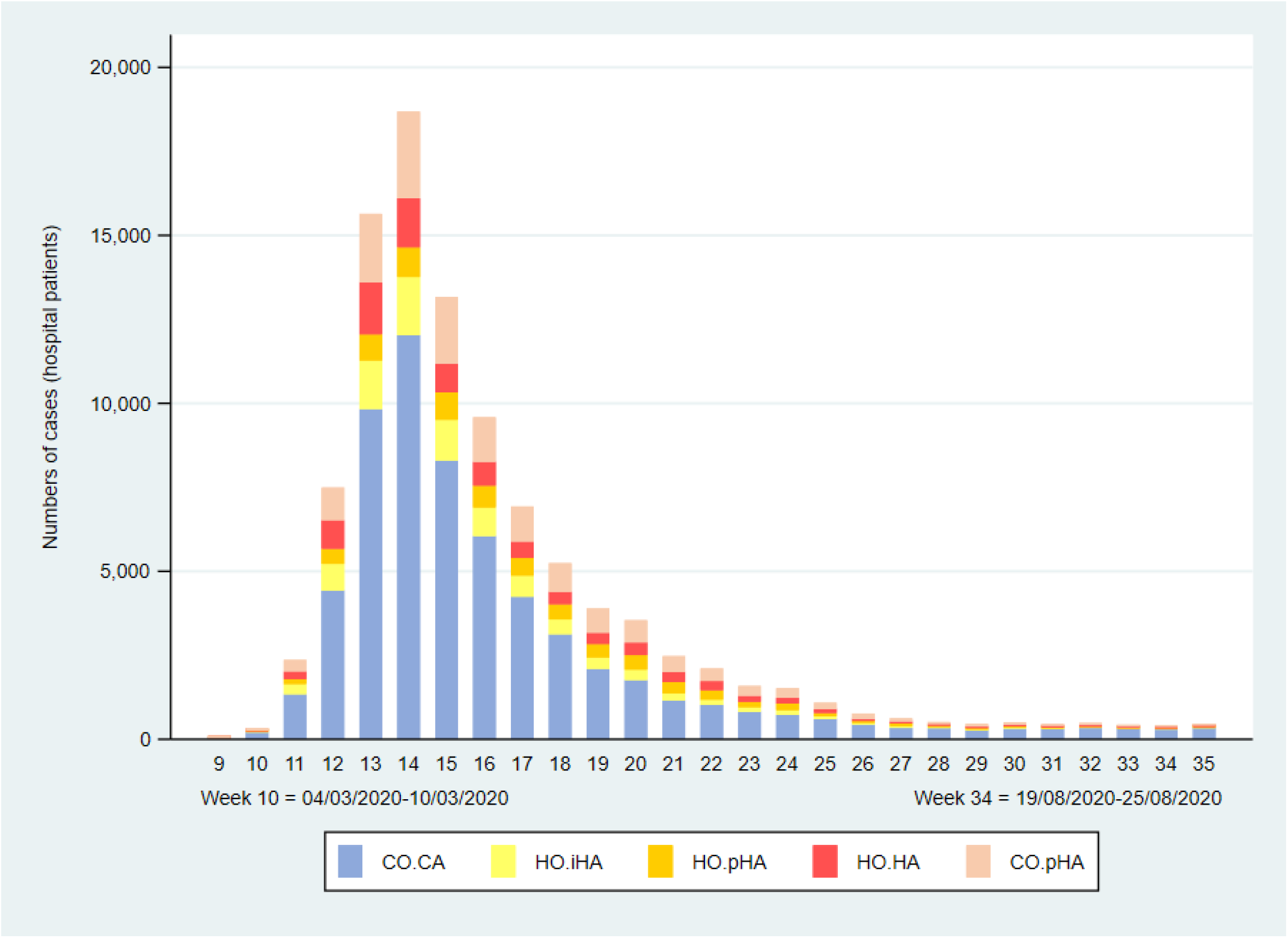
Patients admitted to hospitals in England who tested positive for SARS-CoV-2, showing the weekly numbers of cases classified as hospital-onset indeterminate, probable and definite healthcare-associated (HO.iHA, HO.pHA, HO.HA), community-onset community-acquired (CO.CA) and community-onset possible healthcare-associated (CO.pHA)

**Figure 3:**
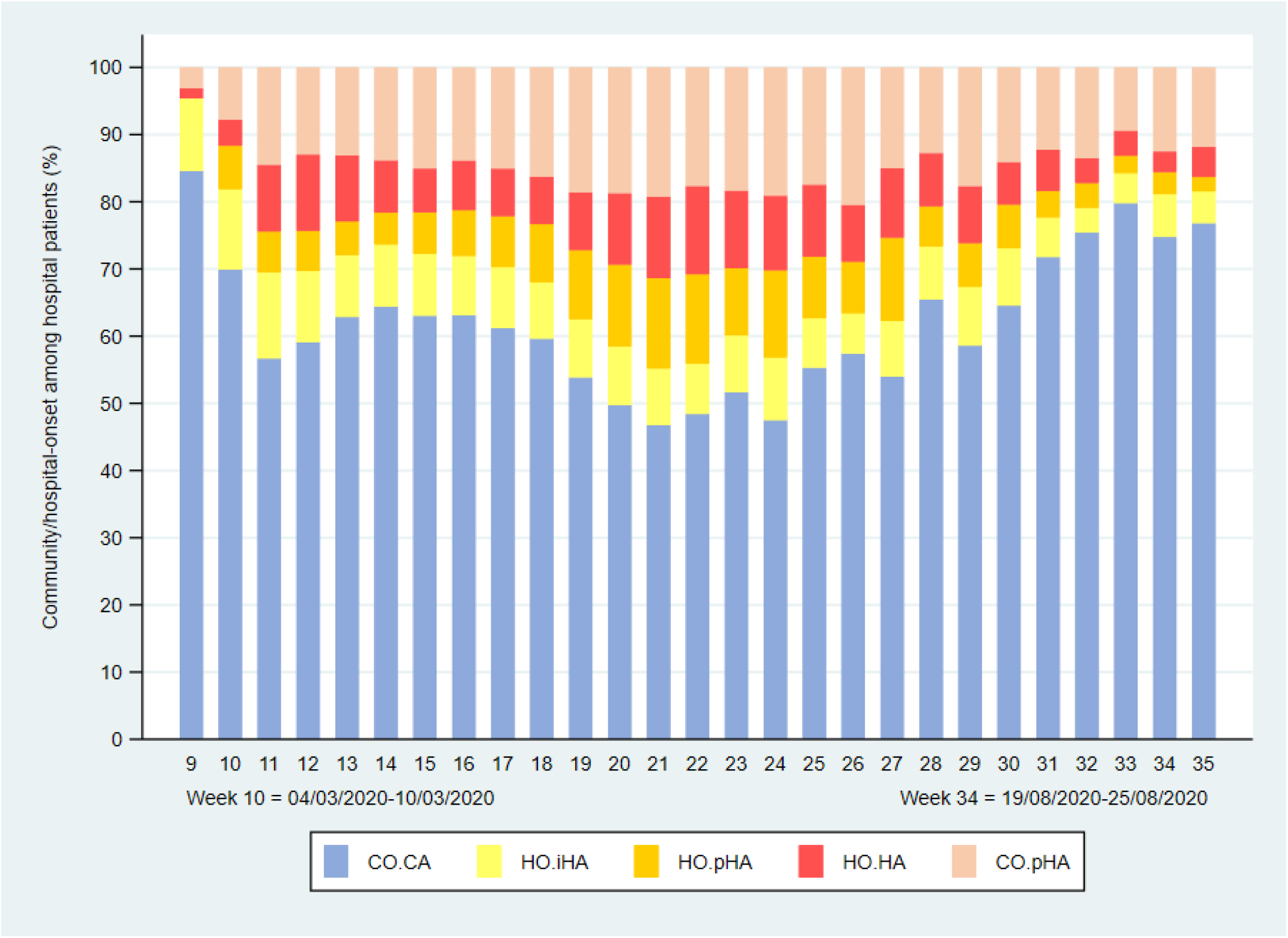
Patients admitted to hospitals in England who tested positive for SARS-CoV-2, showing the weekly percentages of cases classified as hospital-onset indeterminate, probable and definite healthcare-associated (HO.iHA, HO.pHA, HO.HA), community-onset community-acquired (CO.CA) and community-onset possible healthcare-associated (CO.pHA)

There was considerable variation across regions of England in the proportions of hospital patients classified as HO.pHA and HO.HA, from 11.2% (2,427/21,770) in London to 19.3% (3,173/16,427) in the North West NHS region (**Table 2**). A higher proportion of laboratory-confirmed cases linked to Mental Health and Learning Disability NHS Trusts were classified as probable or definite healthcare-associated (54.2%, 1,253/2,310) compared with NHS Acute Trusts (14.3%, 13,875/97,372) (**Table 3**).

**Table 2:** Community-onset and hospital-onset COVID-19 as proportions of all hospital patients who tested positive for SARS-CoV-2 by NHS region.

| **NHS region** | **Hospital patients positive for SARS-CoV-2** | **Community-onset community-acquired** | **Hospital-onset indeterminate healthcare-associated** | **Hospital-onset probable healthcare-associated** | **Hospital-onset definite healthcare-associated** | **Community-onset possible healthcare-associated** |
| --- | --- | --- | --- | --- | --- | --- |
| **London** | 21,770 | 14,296 (65.7%) | 2,089 (9.6%) | 956 (4.4%) | 1,471 (6.8%) | 2,958 (13.6%) |
| **Midlands** | 19,021 | 11,098 (58.4%) | 1,823 (9.6%) | 1,509 (7.9%) | 1,652 (8.7%) | 2,939 (15.5%) |
| **North West** | 16,427 | 9,279 (56.5%) | 1,397 (8.5%) | 1,380 (8.4%) | 1,793 (10.9%) | 2,578 (15.7%) |
| **North East & Yorkshire** | 15,208 | 9,581 (63.0%) | 1,182 (7.8%) | 920 (6.1%) | 1,139 (7.5%) | 2,386 (15.7%) |
| **South East** | 12,873 | 7,659 (59.5%) | 1,153 (9.0%) | 1,040 (8.1%) | 1,247 (9.7%) | 1,774 (13.8%) |
| **East of England** | 10,738 | 6,391 (59.5%) | 1,115 (10.4%) | 785 (7.3%) | 797 (7.4%) | 1,650 (15.4%) |
| **South West** | 4,817 | 2,928 (60.8%) | 386 (8.0%) | 350 (7.3%) | 525 (10.9%) | 628 (13.0%) |
| **Overall** | 100,854<sup>†</sup> | 61,232 (60.7%) | 9,145 (9.1%) | 6,940 (6.9%) | 8,624 (8.6%) | 14,913 (14.8%) |

<sup>†</sup> NHS region was not recorded for 5 CO.CA cases

**Table 3:** Community-onset and hospital-onset COVID-19 as proportions of all hospital patients who tested positive for SARS-CoV-2 by type of provider.

| Healthcare provider | Hospital patients positive for SARS-CoV-2 | Community-onset community-acquired | Hospital-onset indeterminate healthcare-associated | Hospital-onset probable healthcare-associated | Hospital-onset definite healthcare-associated | Community-onset possible healthcare-associated |
| --- | --- | --- | --- | --- | --- | --- |
| NHS Acute Trust | 97,372 | 60,233 (61.9%) | 8,679 (8.9%) | 6,435 (6.6%) | 7,440 (7.6%) | 14,585 (15.0%) |
| Independent (non-NHS providers) | 490 | 303 (61.8%) | 45 (9.2%) | 48 (9.8%) | 65 (13.3%) | 29 (5.9%) |
| NHS Community Trust | 663 | 157 (23.7%) | 121 (18.3%) | 136 (20.5%) | 170 (25.6%) | 79 (11.9%) |
| NHS Mental Health & Learning Disability Trust | 2,310 | 539 (23.3%) | 299 (12.9%) | 320 (13.9%) | 933 (40.4%) | 219 (9.5%) |
| **Overall** | 100,854<sup>†</sup> | 61,232 (60.7%) | 9,145 (9.1%) | 6,940 (6.9%) | 8,624 (8.6%) | 14,913 (14.8%) |

<sup>†</sup> Healthcare provider was not recorded for 5 CO.CA cases

### Characteristics and outcomes of community-onset and hospital-onset COVID-19 cases

The median (IQR) age of hospital patients with a positive test in NHS Acute Trusts was 71 (54-83) years compared with 77 (62-85) years for NHS Mental Health and Learning Disability Trusts. Among NHS Acute Trust hospital patients who tested positive for SARS-CoV-2, older patients (age ≥60 years) were more likely to have a hospital-onset probable or definite healthcare-associated infection (18.5% (12,106/65,534)) than patients under 60 years of age (5.6% (1,769/31,830)) (**Table 4, Figure 4**). Among NHS Mental Health and Learning Disability Trust patients, 55.9% (989/1,769) of laboratory-confirmed COVID-19 cases in patients aged 60 years and older were hospital-onset probable or definite healthcare-associated compared with 48.8% (264/541) of laboratory-confirmed cases in patients under 60 years of age (**Table S1**).

**Table 4:** Characteristics and outcomes of community-onset and hospital-onset laboratory-confirmed COVID-19 cases in NHS Acute Trusts.

|  |  | Hospital patients<br>positive for<br>SARS-CoV-2 | Community-onset<br>community-acquired | Hospital-onset<br>indeterminate<br>healthcare-associated | Hospital-onset<br>probable<br>healthcare-associated | Hospital-onset<br>definite<br>healthcare-associated | Community-onset<br>possible<br>healthcare-associated |
| --- | --- | --- | --- | --- | --- | --- | --- |
|  |  | N=97,372 | n=60,233 | n=8,679 | n=6,435 | n=7,440 | n=14,585 |
| **Age (years)** | <18 years | 1,305 (1.3%) | 901 (1.5%) | 81 (0.9%) | 15 (0.2%) | 66 (0.9%) | 242 (1.7%) |
|  | 18-29 years | 3,714 (3.8%) | 2,957 (4.9%) | 154 (1.8%) | 41 (0.6%) | 94 (1.3%) | 468 (3.2%) |
|  | 30-39 years | 5,769 (5.9%) | 4,589 (7.6%) | 282 (3.3%) | 88 (1.4%) | 108 (1.5%) | 702 (4.8%) |
|  | 40-49 years | 8,263 (8.5%) | 6,415 (10.7%) | 496 (5.7%) | 179 (2.8%) | 238 (3.2%) | 935 (6.4%) |
|  | 50-59 years | 12,779 (13.1%) | 9,316 (15.5%) | 975 (11.2%) | 398 (6.2%) | 542 (7.3%) | 1,548 (10.6%) |
|  | 60-69 years | 13,737 (14.1%) | 8,775 (14.6%) | 1,294 (14.9%) | 731 (11.4%) | 925 (12.4%) | 2,012 (13.8%) |
|  | 70-79 years | 19,151 (19.7%) | 10,370 (17.2%) | 1,964 (22.6%) | 1,537 (23.9%) | 1,797 (24.2%) | 3,483 (23.9%) |
|  | 80+ years | 32,646 (33.5%) | 16,904 (28.1%) | 3,433 (39.6%) | 3,446 (53.6%) | 3,670 (49.3%) | 5,193 (35.6%) |
| **Sex** | Female | 45,121 (46.4%) | 28,106 (46.7%) | 3,806 (43.9%) | 3,050 (47.4%) | 3,526 (47.4%) | 6,633 (45.5%) |
|  | Male | 52,228 (53.6%) | 32,104 (53.3%) | 4,873 (56.1%) | 3,385 (52.6%) | 3,914 (52.6%) | 7,952 (54.5%) |
| **Ethnicity (BAME = black and ethnic minority)** | White | 75,370 (78.4%) | 43,956 (74.2%) | 7,071 (82.1%) | 5,867 (91.7%) | 6,664 (90.3%) | 11,812 (81.7%) |
|  | BAME | 20,762 (21.6%) | 15,325 (25.8%) | 1,546 (17.9%) | 529 (8.3%) | 716 (9.7%) | 2,646 (18.3%) |
| **Charlson score (range 0-16)** | 0 | 21,552 (25.7%) | 15,053 (31.4%) | 1,880 (22.3%) | 815 (13.1%) | 790 (11.3%) | 3,014 (21.3%) |
|  | 1 | 20,574 (24.6%) | 13,002 (27.1%) | 2,100 (24.9%) | 1,217 (19.5%) | 1,309 (18.7%) | 2,946 (20.8%) |
|  | 2+ | 41,641 (49.7%) | 19,913 (41.5%) | 4,466 (52.9%) | 4,199 (67.4%) | 4,888 (70.0%) | 8,175 (57.8%) |
| **Length of stay (hospital spell)** | median (IQR) days | - | 5 (1-11) | 11 (7-19) | 19 (14-28) | 41 (28-72) | 6 (3-13) |
| **Length of stay (post-test)** | median (IQR) days | - | 4 (1-10) | 7 (3-15) | 9 (4-18) | 13 (6-27) | 6 (3-13) |
| **Mortality (28 days post-test)** |  | 29,073 (29.9%) | 15,620 (25.9%) | 3,003 (34.6%) | 2,840 (44.1%) | 2,886 (38.8%) | 4,724 (32.4%) |

**Figure 4:**
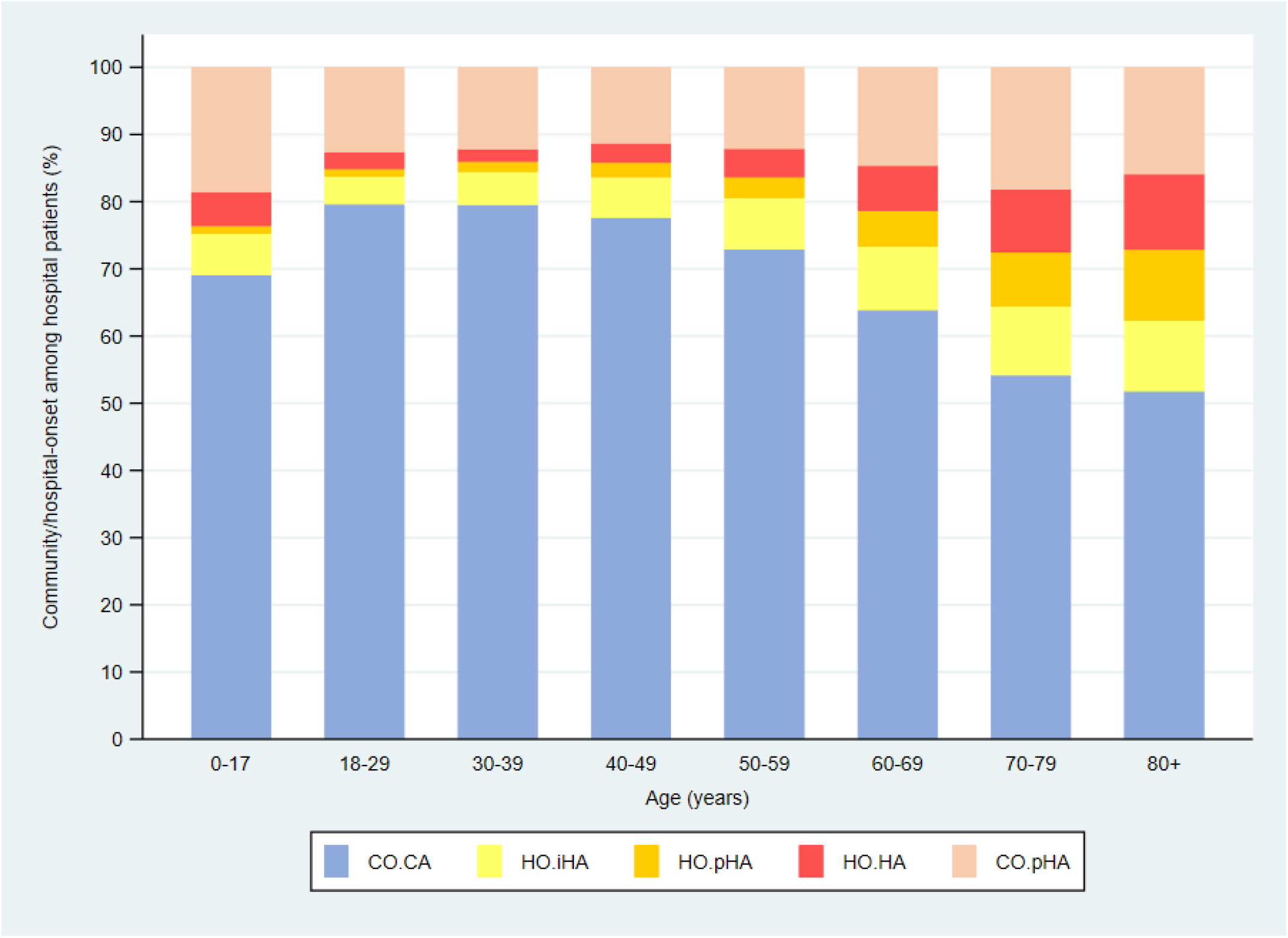
Patients admitted to NHS Acute Trust hospitals in England who tested positive for SARS-CoV-2, showing the percentages of cases classified as hospital-onset indeterminate, probable and definite healthcare-associated (HO.iHA, HO.pHA, HO.HA), community-onset community-acquired (CO.CA) and community-onset possible healthcare-associated (CO.pHA) by age group

Among patients in Acute Trusts, HO.HA cases had the longest total length of stay (median 41, IQR 28-72 days) and longest post-test length of stay (median 13, IQR 6-27 days (**Table 4**). In Mental Health and Learning Disability Trusts, the median total length of stay for HO.HA cases was 83 days (IQR 44-231 days); the median post-test length of stay for these cases was 29 (IQR 12-79) days (**Table S1**).

The proportions of patients with 2 or more comorbidities (Charlson index ≥2) in NHS Acute Trust ranged from 42% in CO.CA cases to 70% in HO.HA cases (**Table 4**). HO.pHA and HO.HA patients in NHS Acute Trusts had 41.3% (5,726/13,875) 28-day COVID-related mortality, compared with 25.9% (15,620/60,233) in CO.CA cases (**Table 4, Table S2**). In patients in NHS Mental Health and Learning Disability Trusts, 28-day mortality among HO.pHA and HO.HA cases was 21.9% (274/1,253) (**Table S1**).

## Discussion

Our study of healthcare-associated COVID-19 in hospital patients, encompassing the first phase of the COVID-19 pandemic in England, is the first to use large-scale national data. We found that 15% of patients admitted with or diagnosed during admission with SARS-CoV-2 infection during the first 6 months of the pandemic were hospital-onset probable or definite healthcare-associated, representing 5% of all laboratory-confirmed COVID-19 cases during this period. A further 15% of laboratory-confirmed cases in hospital patients who had COVID-19 were possibly healthcare-associated but with a first positive test after discharge.

Our results are descriptive. We did not attempt more in-depth analyses, our aim being to present an overall picture of healthcare-associated COVID-19. Further analyses of national data might be useful, although the time-varying nature of many of the factors involved in COVID-19, particularly testing practices, and fundamental differences between community and hospital populations and between hospitals may preclude a meaningful analysis of such observational data. Instead, smaller prospective studies with well-characterized patient cohorts and complete epidemiological data may be more useful in determining risk factors for healthcare-associated SARS-CoV-2 infection and providing evidence to inform infection prevention and control measures in healthcare settings.

### In the context of other studies and reports

Our estimated proportion for HO.pHA and HO.HA combined was consistent with other reports: a study in 10 UK hospitals and 1 Italian hospital reported 13% up to 28^th^ April,^4^ and a single London hospital reported 15% between 2^nd^ March and 12^th^ April.^5^ Although our data covered the period up to 30^th^ August, 75% of laboratory-confirmed cases linked to hospital records occurred during March and April, and the HO.pHA and HO.HA proportion in our dataset for those two months (14%) was only slightly lower than our estimate for the whole 6-month period. Hospital-onset cases to 30^th^ August represented 6.4% of all laboratory-confirmed COVID-19 cases in Scotland and 10.5% of all laboratory-confirmed COVID-19 cases in Wales.^13,14^ The lower proportion (5.3%) in England may reflect differences in hospital admissions or testing over the peak months.

There is a growing international literature on nosocomial SARS-CoV-2 infection, from single ward or department reports,^15-23^ to hospital-wide studies,^24-26^ but only one (from Malta) based on limited national surveillance data.^27^ The lull between phases of the pandemic in Europe has allowed prospective studies to be planned, but these have yet to report.^28^ Early estimates for nosocomial COVID-19 will be highly variable because responses to the pandemic changed rapidly over time, most notably in SARS-CoV-2 testing in the community and in healthcare settings.

The 28-day mortality rate (26%) for community-onset community-acquired cases admitted to NHS Acute hospitals in our study was consistent with in-hospital mortality reported directly by NHS Acute hospitals (26%).^29^ Hospital-onset cases experienced higher mortality, as expected given their higher median age (79 years for HO.HA cases compared with 66 years for CO.CA cases) and pre-existing conditions. The higher proportion of hospital-onset cases in Mental Health and Learning Disability Trusts probably reflects longer stays in these settings, which include residential and secure psychiatric units, compared with Acute Trusts; COVID-19 mortality among hospital-onset cases in Mental Health and Learning Disability Trust patients (22% for HO.pHA and HO.HA) was lower than in community-onset cases in acute hospitals. However, as noted above, these crude comparisons do not consider a multiplicity of differences between patient groups. Mental health hospitals had relatively fewer inpatients with COVID-19, therefore nosocomial proportions are based on much smaller denominators. It is also possible that case detection may have been sub-optimal in such settings.

### Strengths and limitations

The strength of our study is that centralised, routinely collected national data sources were used which recorded all positive COVID-19 test results from Pillars 1 and 2 of the UK government’s public testing programmes, and all NHS hospital attendances and admissions in England. The latter represented approximately 98% of all hospital activity in the country.^30^ Pillar 1 tests were provided by NHS and PHE laboratories for community cases from January 16^th^ to March 12^th^, 2020, for hospitalised cases and the investigation of care home outbreaks from March 12^th^ to March 31^st^, 2020, and for healthcare workers and their families from April 1^st^ onwards (where additional capacity was available). Pillar 2 testing was delivered by central government through academic, public and private partnerships: from April 1^st^ 2020 it was progressively rolled out to key workers in the NHS, social care and other critical sectors; from May 23^rd^ the general population could also access testing from this route; from August 1^st^ it was used to test asymptomatic staff and residents in care homes and for asymptomatic contacts in outbreak investigations. We saw a large expected decrease in the proportion of cases with a temporal link to a hospital record (from 79% in March to 7% in August), as testing policy across the UK expanded from an initial focus on testing in hospitals to community testing.

The main limitation of our study is that our case numbers reflect national testing activity, not the true number of cases in the population, and this activity was severely constrained by testing capacity during the phase of the pandemic covered by our study. As of September 8^th^, 2020, it was estimated from household survey data that approximately 2.8 million people (95% CI 2.4 to 3.2 million people) aged 16 years and over would have antibodies to COVID-19.^31^ Therefore, the 15,564 HO.pHA and HO.HA cases during this six-month period represent approximately 1% of all cases in England at the time of this estimate. While numbers of hospital-onset cases should be closer to the true number of cases, assuming that patients in hospital were more likely to be tested, an unknown number of asymptomatic cases will have been missed where inpatients were not routinely swabbed. Systematic testing of inpatients in hospitals in England did not start until 24^th^ June. Similarly, cases classified as probable or definite hospital-onset may have been infected before admission or during the first 7 days of admission but were not tested until they became symptomatic, or they may have had negative test results, which were not available for our analysis. The overall effect of these limitations will likely have been to over-estimate probable and definite hospital-onset case numbers. Linkage of national surveillance and hospital activity data will be imperfect, and our algorithm made assumptions in assigning a priority order when test result data linked to more than one attendance or admission. For CO.pHA cases, we treated all hospital admissions equally regardless of duration of healthcare exposure. Conversely, we would not have captured infections potentially acquired from primary care, outpatient and emergency department attendances. Our data did not allow us to identify healthcare workers whose infection may have been acquired in the workplace, and we would have misclassified these cases as community-acquired. Our analysis used test and admission dates rather than dates and times because time of day was not recorded in our test or inpatient data sources, therefore our results will not be exactly comparable with classifications based on exact time, i.e., ≤48 hours rather than ≤2 days.

### Implications for policy and practice

Frontline healthcare workers were identified as a high-risk group during the first phase of the pandemic,^32^ highlighting an urgent need for personal protective equipment and procedures. This was of particular importance to protect staff for their own safety, to prevent onward transmission to patients, to minimise staff absence at a time of extraordinarily high demand on the NHS.^33^ andbecause patients may be in an incubation period or have a false negative test.^24^ Arguably, countries which had direct experience of SARS-CoV-1 were better prepared to respond to the risk of healthcare-associated COVID-19.^25^ In countries such as the UK, which are experiencing distinct phases of the pandemic, preparedness for subsequent phases should be better than during the first phase, including the availability of comprehensive guidelines for infection prevention and control in different healthcare settings and for care of patients at increased risk of severe COVID-19 illness.^34^ To reduce transmission in hospitals from patient to patient and from patient to healthcare worker, the UK now recommends pre-admission testing of all patients who are to be admitted for elective procedures, testing on admission for emergency admissions, and testing at 3-7 days post-admission. To reduce transmission of asymptomatic or pre-symptomatic COVID-19 from healthcare worker to healthcare worker and from healthcare worker to patient, hospitals are recommended to screen staff on a weekly basis in periods of higher community prevalence, during hospital outbreaks and when cases of nosocomial COVID-19 are detected. More extensive (3 times a week) screening of patient facing healthcare workers in the NHS has recently been rolled out.

Currently, very limited rapid emergency care testing will likely have two main consequences for nosocomial infection: firstly, within A&E, because positive and negative patients cannot be readily identified in a setting of crowded units and waiting areas; secondly, patients with unknown COVID-19 status are admitted to a hospital bed, typically in 4-, 6- or 10-bedded bays. Lastly, and crucially, the proportion of NHS hospital beds in England that are in single rooms is approximately 20%, which severely constrains capacity to prevent airborne virus transmission.

## Data Availability

Requests for aggregate data not otherwise shown in the manuscript and supplementary files can be sent to the corresponding author.

## Acknowledgements

We would like to thank the Public Health England (PHE) National Incident Coordination Centre (NICC) Epidemiology Cell (EpiCell) and the PHE Data Lake team.

---

**Figure S1:**
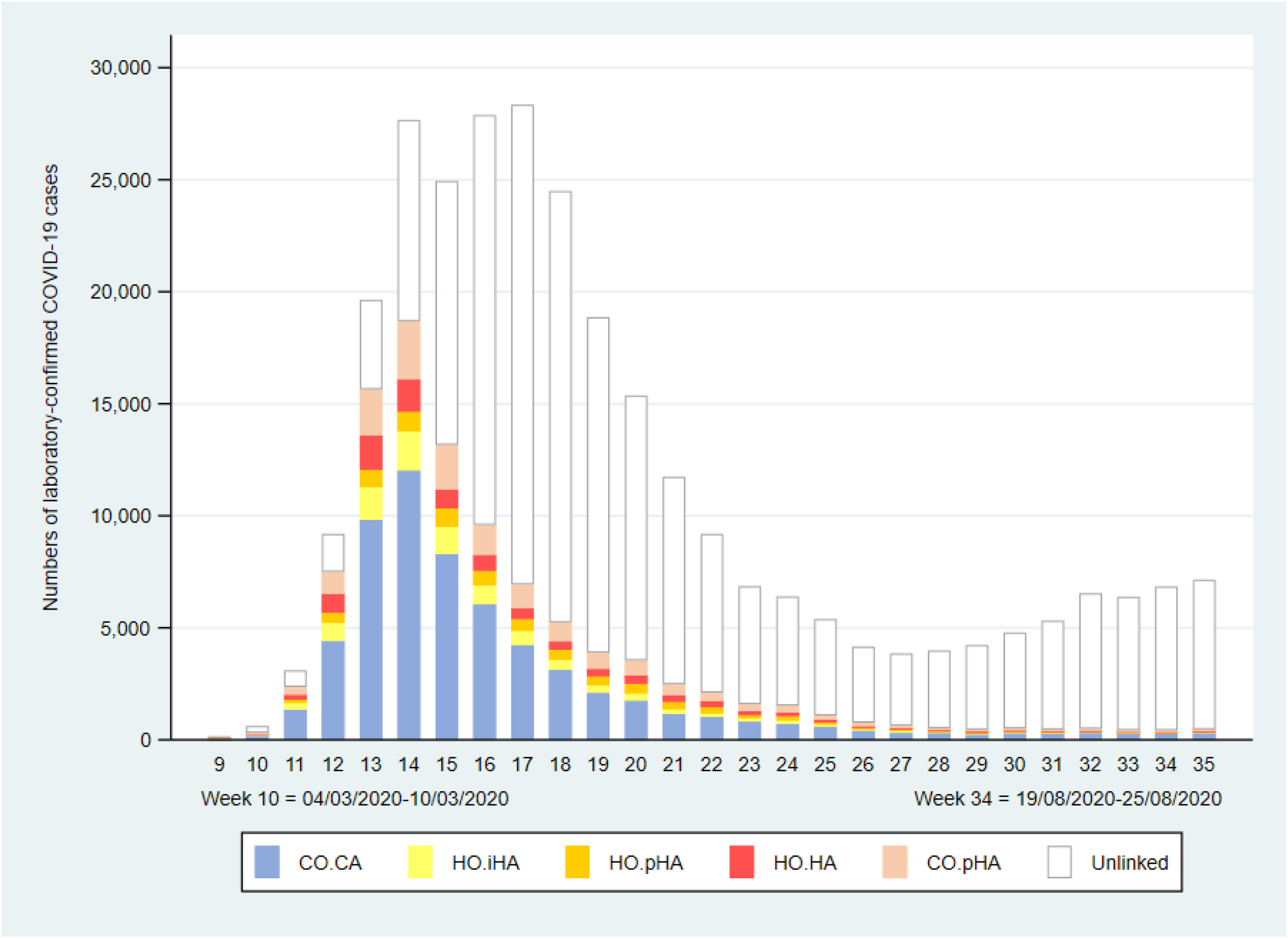
Patients admitted to hospitals in England who tested positive for SARS-CoV-2, showing the weekly numbers of cases classified as hospital-onset indeterminate, probable and definite healthcare-associated (HO.iHA, HO.pHA, HO.HA), community-onset community-acquired (CO.CA) and community-onset possible healthcare-associated (CO.pHA) and laboratory-confirmed COVID-19 cases not linked to a hospital admission (Unlinked)

**Table S1:** Characteristics and outcomes of community-onset and hospital-onset laboratory-confirmed COVID-19 cases in NHS Mental Health & Learning Disability Trusts (March to August 2020)

|  |  | Hospital patients<br>positive for SARS-CoV-2 | Community-onset<br>community-acquired | Hospital-onset<br>indeterminate<br>healthcare-associated | Hospital-onset<br>probable healthcare-associated | Hospital-onset<br>definite healthcare-associated | Community-onset<br>possible healthcare-associated |
| --- | --- | --- | --- | --- | --- | --- | --- |
|  |  | N=2,310 | n=539 | n=199 | n=320 | n=933 | n=219 |
| **Age (years)** | 0-39 years | 229 (9.9%) | 73 (13.5%) | 21 (7.0%) | 21 (6.6%) | 99 (10.6%) | 15 (6.9%) |
|  | 40-49 years | 119 (5.2%) | 48 (8.9%) | 10 (3.3%) | 14 (4.4%) | 41 (4.4%) | 6 (2.7%) |
|  | 50-59 years | 193 (8.4%) | 67 (12.4%) | 20 (6.7%) | 23 (7.2%) | 66 (7.1%) | 17 (7.8%) |
|  | 60-69 years | 269 (11.7%) | 69 (12.8%) | 32 (10.7%) | 31 (9.7%) | 111 (11.9%) | 26 (11.9%) |
|  | 70-79 years | 530 (22.9%) | 111 (20.6%) | 63 (21.1%) | 67 (20.9%) | 244 (26.2%) | 45 (20.6%) |
|  | 80+ years | 970 (42.0%) | 171 (31.7%) | 153 (51.2%) | 164 (51.3%) | 372 (39.9%) | 110 (50.2%) |
| **Sex** | Female | 1,094 (47.4%) | 255 (47.3%) | 152 (50.8%) | 146 (45.6%) | 440 (47.2%) | 101 (46.1%) |
|  | Male | 1,216 (52.6%) | 284 (52.7%) | 147 (49.2%) | 174 (54.4%) | 493 (52.8%) | 118 (53.9%) |
| **Ethnicity** | White | 2,121 (92.6%) | 506 (96.0%) | 276 (92.3%) | 292 (91.8%) | 835 (89.9%) | 212 (97.3%) |
|  | Black & ethnic minority | 170 (7.4%) | 21 (4.0%) | 23 (7.7%) | 26 (8.2%) | 94 (10.1%) | 6 (2.8%) |
| **Charlson score (range 0-16)** | 0 | 918 (41.2%) | 165 (35.6%) | 146 (49.2%) | 130 (40.6%) | 409 (44.0%) | 68 (31.1%) |
|  | 1 | 508 (22.8%) | 121 (26.1%) | 64 (21.6%) | 67 (20.9%) | 219 (23.6%) | 37 (16.9%) |
|  | 2+ | 803 (36.0%) | 177 (38.2%) | 87 (29.3%) | 123 (38.4%) | 302 (32.5%) | 114 (52.0%) |
| **Length of stay (hospital spell)** | median (IQR) days | - | 5 (1-12) | 18 (10-34) | 27 (17-47) | 83 (44-231) | 9 (4-21) |
| **Length of stay (post-test)** | median (IQR) days | - | 5 (1-12) | 13 (5-28) | 16 (6-34) | 29 (12-79) | 9 (4-21) |
| **Mortality (28 days post-test)** |  | 518 (22.4%) | 120 (22.4%) | 75 (25.1%) | 81 (25.3%) | 193 (20.7%) | 49 (22.4%) |

**Table S2:**
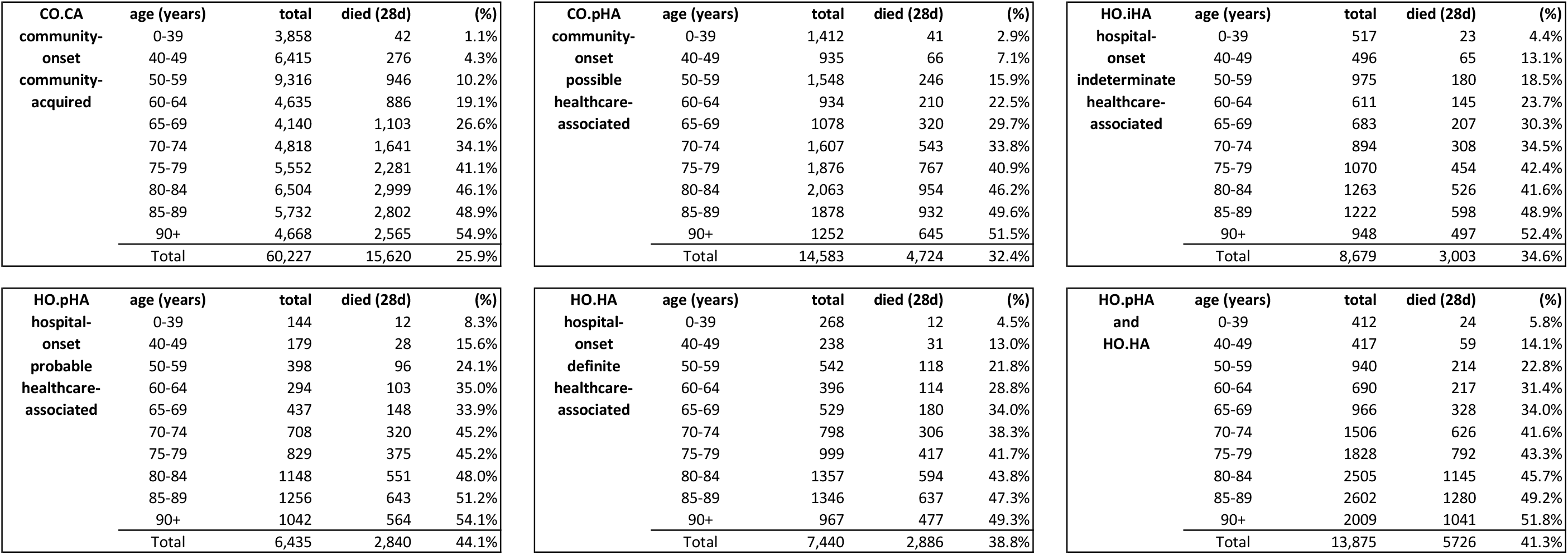
Mortality at 28 days (after first positive SARS-CoV-2 PCR test) by community- or hospital-onset COVID-19 case classification in patients admitted to NHS Acute Trust hospitals (March to August 2020)

## References

1. Zhou Q, Gao Y, Wang X, et al. Nosocomial infections among patients with COVID-19, SARS and MERS: a rapid review and meta-analysis. Ann Transl Med 2020; 8(10): 629.

2. Wang D, Hu B, Hu C, et al. Clinical Characteristics of 138 Hospitalized Patients With 2019 Novel Coronavirus–Infected Pneumonia in Wuhan, China. JAMA 2020; 323(11): 1061–9.

3. DELVE Initiative. DELVE Scoping report on hospital and health care acquisition of COVID-19 and its control. London: The Royal Society, 2020. https://rs-delve.github.io/reports/2020/07/06/nosocomial-scoping-report.html

4. Carter B, Collins JT, Barlow-Pay F, et al. Nosocomial COVID-19 infection: examining the risk of mortality. The COPE-Nosocomial Study (COVID in Older PEople). J Hosp Infect 2020; 106(2): 376–84.

5. Rickman HM, Rampling T, Shaw K, et al. Nosocomial transmission of COVID-19: a retrospective study of 66 hospital-acquired cases in a London teaching hospital. Clin Infect Dis 2020.

6. Department of Health and Social Care (DHSC). Coronavirus (COVID-19): Scaling up our testing programmes. London: DHSC, 2020. https://assets.publishing.service.gov.uk/government/uploads/system/uploads/attachment_data/file/878121/coronavirus-covid-19-testing-strategy.pdf

7. Public Health England (PHE). Laboratory reporting to Public Health England: A guide for diagnostic laboratories. London: PHE, 2016. https://assets.publishing.service.gov.uk/government/uploads/system/uploads/attachment_data/file/739854/PHE_Laboratory_Reporting_Guidelines.pdf

8. NHS Digital. Secondary Uses Service (SUS). https://digital.nhs.uk/services/secondary-uses-service-sus (accessed 18/10/2020).

9. Health and Social Care Information Centre (HSCIC). Methodology to create provider and CIP spells from HES APC data 2014. http://content.digital.nhs.uk/media/11859/provider-spells-methodology/pdf/spells_methodology.pdf/

10. Quan H, Sundararajan V, Halfon P, et al. Coding algorithms for defining comorbidities in ICD-9-CM and ICD-10 administrative data. Med Care 2005; 43(11): 1130–9.

11. Public Health England (PHE). PHE data series on deaths in people with COVID-19: Technical summary - 12 August update. London: PHE, 2020.https://assets.publishing.service.gov.uk/government/uploads/system/uploads/attachment_data/file/916035/RA_Technical_Summary_-_PHE_Data_Series_COVID_19_Deaths_20200812.pdf

12. European Centre for Disease Prevention and Control (ECDC). Surveillance definitions for COVID-19. https://www.ecdc.europa.eu/en/covid-19/surveillance/surveillance-definitions (accessed 02/12/2020).

13. Health Protection Scotland (HPS). Hospital onset COVID-19 cases in Scotland: Week ending 1 March to week ending 30 August 2020. Glasgow: HPS, 2020. https://beta.isdscotland.org/find-publications-and-data/population-health/covid-19/hospital-onset-covid-19-cases-in-scotland/23-september-2020/

14. Public Health Wales (Iechyd Cyhoeddus Cymru). https://public.tableau.com/profile/public.health.wales.health.protection#!/vizhome/RapidCOVID-19virology-Public/Headlinesummary.

15. Bays DJ, Nguyen MH, Cohen SH, et al. Investigation of Nosocomial SARS-CoV-2 Transmission from Two Patients to Health Care Workers Identifies Close Contact but not Airborne Transmission Events. Infect Control Hosp Epidemiol 2020: 1–22.

16. Hara T, Yamamoto C, Sawada R, et al. Infection risk in a gastroenterological ward during a nosocomial COVID-19 infection event. J Med Virol 2020.

17. Heng AL, Ooi CC, Wen Eu BJ, San Kiew Y, Wong ASK, Da Zhuang K. The bug stops with me: Prevention of COVID-19 nosocomial transmission during radiographic procedures. J Med Imaging Radiat Sci 2020.

18. Jewkes SV, Zhang Y, Nicholl DJ. Nosocomial spread of COVID-19: lessons learned from an audit on a stroke/neurology ward in a UK district general hospital. Clin Med (Lond) 2020; 20(5): e173–e7.

19. Jung J, Hong MJ, Kim EO, Lee J, Kim MN, Kim SH. Investigation of a nosocomial outbreak of coronavirus disease 2019 in a paediatric ward in South Korea: successful control by early detection and extensive contact tracing with testing. Clin Microbiol Infect 2020.

20. Lakhani K, Minguell J, Guerra-Farfan E, et al. Nosocomial infection with SARS-CoV-2 and main outcomes after surgery within an orthopaedic surgery department in a tertiary trauma centre in Spain. Int Orthop 2020: 1–9.

21. Biernat MM, Zinczuk A, Biernat P, et al. Nosocomial outbreak of SARS-CoV-2 infection in a haematological unit - High mortality rate in infected patients with haematologic malignancies. J Clin Virol 2020; 130: 104574.

22. Sánchez MD, Sánchez M, De La Morena JM, et al. Nosocomial SARS-CoV-2 infection in urology departments: Results of a prospective multicentric study. Int J Urol 2020.

23. Ingels A, Bibas S, Da Costa JB, et al. Surgery and COVID-19: Balancing the nosocomial risk a french academic center experience during the epidemic peak. Br J Surg 2020.

24. Correa-Martinez CL, Schwierzeck V, Mellmann A, Hennies M, Kampmeier S. Healthcare-Associated SARS-CoV-2 Transmission-Experiences from a German University Hospital. Microorganisms 2020; 8(9).

25. Kim SW, Jo SJ, Lee H, et al. Containment of a healthcare-associated COVID-19 outbreak in a university hospital in Seoul, Korea: A single-center experience. PLoS One 2020; 15(8): e0237692.

26. Khonyongwa K, Taori SK, Soares A, et al. Incidence and outcomes of healthcare-associated COVID-19 infections: significance of delayed diagnosis and correlation with staff absence. J Hosp Infect 2020.

27. Micallef S, Piscopo TV, Casha R, et al. The first wave of COVID-19 in Malta; a national cross-sectional study. PLoS One 2020; 15(10): e0239389.

28. Saadatian-Elahi M, Picot V, Hénaff L, et al. Protocol for a prospective, observational, hospital-based multicentre study of nosocomial SARS-CoV-2 transmission: NOSO-COR Project. BMJ Open 2020; 10(10): e039088.

29. Docherty AB, Harrison EM, Green CA, et al. Features of 20 133 UK patients in hospital with covid-19 using the ISARIC WHO Clinical Characterisation Protocol: prospective observational cohort study. BMJ 2020; 369: m1985.

30. Herbert A, Wijlaars L, Zylbersztejn A, Cromwell D, Hardelid P. Data Resource Profile: Hospital Episode Statistics Admitted Patient Care (HES APC). Int J Epidemiol 2017; 46(4): 1093–i.

31. Office for National Statistics (ONS). Coronavirus (COVID-19) Infection Survey pilot: England, Wales and Northern Ireland, 25 September 2020. Newport, 2020. https://www.ons.gov.uk/peoplepopulationandcommunity/healthandsocialcare/conditionsanddiseases/bulletins/coronaviruscovid19infectionsurveypilot/englandwalesandnorthernireland25september2020#antibody-data-for-england

32. Nguyen LH, Drew DA, Graham MS, et al. Risk of COVID-19 among front-line health-care workers and the general community: a prospective cohort study. Lancet Public Health 2020; 5(9): e475–e83.

33. Khonyongwa K, Taori SK, Soares A, et al. Incidence and outcomes of healthcare-associated COVID-19 infections: significance of delayed diagnosis and correlation with staff absence. J Hosp Infect 2020.

34. Public Health England (PHE). COVID-19: infection prevention and control (IPC). https://www.gov.uk/government/publications/wuhan-novel-coronavirus-infection-prevention-and-control (accessed 05/11/2020).

